# A Healthy Buildings Guideline for the COVID-19 Pandemic and Beyond

**DOI:** 10.1101/2020.11.30.20241406

**Authors:** Clifford Federspiel

## Abstract

Public health experts have confirmed that airborne transmission of SARS-CoV-2 (COVID-19) is one of the primary mechanisms of infection (CDC, 2020). In addition to social distancing, mask wearing and hand washing, experts now recommend increasing the ventilation and filtration of indoor air. While there is widespread consensus on this general approach, to date there are no published guidelines for the levels of ventilation, filtration, etc. that are required to control the pandemic. This is an urgent concern because colder weather in the Northern Hemisphere has moved social activity indoors where the risk of infection is higher.

---

We propose a Guideline that provides a Criterion for integrating the effects of engineering and administrative controls with personal protective equipment (PPE) for indoor environments. The Guideline takes into account ventilation, filtration, temperature control, humidity control, masks, occupant density, occupancy category and activity. The design of the Guideline integrates recently published research regarding COVID-19 characteristics (a topic of ongoing scientific investigation) with well-established models for contaminant accumulation and infection risk (Wells-Riley), and is informed by the SIR model of epidemic dynamics. We mathematically determine a minimum threshold for the loss rate (combination of air change rate, removal rate by filtration, inactivation rate, and settling rate) that will keep the expected number of secondary infections from a single infected person less than 1.0 over the sequence of activities performed by the infected person while they are infectious. If the expected number of secondary infections is less than 1.0, then the number of infections at the population level will decrease.

We show how the Guideline can be used in conjunction with existing tabulated air quality standards. We also illustrate the importance of masks and occupant density. Though the Guideline has been developed with SARS-CoV-2 in mind, it could also be applied to future epidemics and other pathogens using different pathogen-specific characteristics.

## Single Zone Model of Well-Mixed Indoor Air with Recirculation

Using a control volume that includes the indoor space, but does not include air-handling equipment, pathogen accumulation indoors is modeled with the following equation.

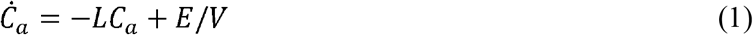

The loss rate, *L*, has three terms.

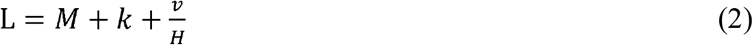

The first term, *M*, is the mechanical air change rate. It includes the effects of ventilation, recirculation, filtration, and inactivation by permanent and temporary air-handling equipment.

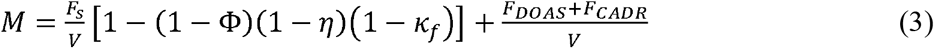

The second term in Equation 2, *k*, is the natural inactivation rate of a pathogen. A model for inactivation of aerosols containing SARS-CoV-2 as a function of temperature, relative humidity, and UV index can be found at DHS (2020). The DHS model predicts that at 22 degC, 20% relative humidity, and UV index = 0, the inactivation rate of SARS-CoV-2 is zero. Increasing the relative humidity to 40% increases the inactivation rate to 0.71/hour. Increasing the temperature to 30 degC while keeping the relative humidity at 20% results in an inactivation rate of 0.63/hour.

The third term in Equation 2,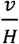, is the droplet settling rate. The settling velocity, *v*, can be modeled with curve fits to experimental data. Droplets shrink due to evaporation after being emitted. The equilibrium diameter can be determined using Kohler theory, with an equation such as the one described by Lewis (2008).

If a pathogen is released into the indoor air at a quasi-steady rate, then the steady-state concentration of the pathogen indoors is as follows

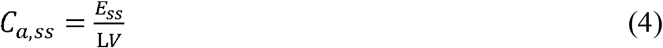

So, the larger the value of *L*, the smaller the value of the steady-state pathogen concentration. Also, the larger the volume of the space, V, the smaller the value of the steady-state pathogen concentration.

## Impact of Wearing Masks

If the emitter and susceptible occupants are wearing masks, then the steady-state concentration in the inhaled breath of the susceptible occupants is as follows

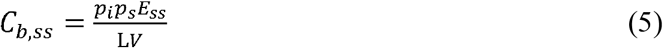

where *p*_*i*_ is the penetration ratio of the mask worn by an infectious emitter and *p*_*s*_ is the penetration ratio of a mask worn by a susceptible occupant. The penetration ratio of a filter is the ratio of the downstream concentration to the upstream concentration. Mathematically, the penetration ratio is equal to one minus the efficiency. The penetration ratio of a mask worn by an infectious emitter might differ from the penetration ratio of a mask worn by a susceptible occupant even if they wear nominally identical masks because the emitted droplets are larger and contain more water than inhaled droplets.

The following table shows filtration efficiency and penetration ratios for infected and susceptible occupants (from Jimenez 2020). The efficiency and penetration ratio for surgical masks worn by susceptible occupants are derived from the results of Oberg and Brosseau (2008) for dental masks after using the same discount used by Jimenez (2020) for surgical masks worn by infected occupants.

## Wells-Riley Infection Model

The Wells-Riley model of infection risk uses an exponential distribution for the probability of infection as a function of the dose. Wells-Riley uses units of quanta to represent the number of virions. One quantum is the dose corresponding to a 63% probability of infection. When concentration is expressed in units of quanta per unit volume and emission is expressed in units of quanta per unit time, the probability of a single person being infected when exposed to air with a constant quanta concentration is as follows

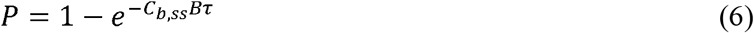

where *C*_*b,ss*_ is the quanta per unit volume, B is the breathing rate of a susceptible person, and τ is the time duration of the activity (exposure duration). If there are S susceptible occupants involved in an activity, then the expected number of people infected is SP.

There is limited information about quanta emission rates for SARS-CoV-2. But there have been a small number of studies that have estimated *E*_*ss*_ under some conditions. Miller et al. (2020) estimated *E*_*ss*_ = 970 quanta/hour for the Skagit Valley Chorale superspreading event using WR. Buanano et al. (2020) estimated a range of values of *E*_*ss*_ for SARS-CoV-2 based on the range of concentrations found in respiratory fluid of infected people and the estimated infectivity ratio of SARS-CoV-1. Bazant and Bush (2020) estimated the quanta concentration in exhaled breath for a range of expiratory types. The following table lists quanta concentration in exhaled breath for some of the expiratory types listed in Bazant and Bush (2020).

For the Guideline, we use a weighted average of expiratory type concentrations along with an appropriate breathing rate to determine *E*_*ss*_. We also assume that there is a single emitter. While there have been some documented examples of multiple emitters in a single activity, the fraction of the population that is infected is low enough that we feel it is acceptable to neglect situations involving more than one infected occupant for the purpose of creating a Guideline.

## SIR Model

The SIR model of the dynamics of epidemics has three state variables, commonly referred to as compartments. The three compartments represent the people in a population who are susceptible to infection (S), the people who are infected (I), and the people who either recovered or who have died (R). The differential equations for the SIR model are as follows.

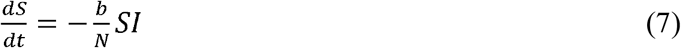

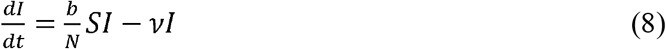

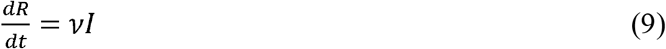

where N = S + I + R is the total number of people in the population, b is the effective contact rate, bS/N is the mean infection rate and ν is the mean recovery rate.

The SIR model reflects the fact that in epidemics, the number of infections will increase when the mean infection rate is greater than the mean recovery rate. Conversely, the number of infections will decrease when the mean infection rate is less than the mean recovery rate. The reproduction number is the mean infection rate divided by the mean recovery rate. At the beginning of an epidemic, N = S, and the initial reproduction number is b/ν. The reproduction number is the average number of secondary infections from each infected person. The initial reproduction number is the average number of secondary infections from the initial infected person when all members of the population are susceptible. We use this interpretation of the reproduction number and the fact that a value less than 1.0 causes the number of infections to decrease as an objective of the Guideline.

## Proposed COVID-19 Indoor Guideline

The principle behind the Guideline is to choose engineering and administrative controls for indoor environments (e.g., ventilation, filtration, temperature control, humidity control, occupant density, masks, etc.) that will result in a reproduction number less than 1.0 so that an epidemic is brought under control. The objective of the Guideline is not individual risk, but instead infection control at the population level. Bazant and Bush (2020) developed an indoor reproduction number for a single discrete activity over a time period, *τ*. We propose a Criterion that is based on keeping the expected number of secondary infections from an infected person less than 1.0 over the sequence of activities the infected person performs over the entire duration that the infected person is infectious. This expected number of secondary infections is equivalent to the reproduction number of the SIR model.

An infected person will engage in a sequence of activities (e.g., commute to work, work morning shift, go out to lunch, work afternoon shift, commute home, etc.) over a period of days while they are infectious. For example, the following calendar shows a sequence of 68 activities performed by a fictitious person who becomes infected in the 9^th^ hour of the first day of the calendar. The onset of symptoms begins in the 9^th^ hour of the fifth day, and this person first becomes infectious in the 9^th^ hour of the third day. The light gray time periods denote the incubation period prior to becoming infectious. The light orange time periods denote the pre-symptomatic period when the person is infectious but not yet showing symptoms. The darker orange shows the time periods when the person is exhibiting symptoms and is infectious, and the darker gray periods show the time after the person is no longer infectious. During the lighter and darker orange periods this person engages in activities ranging from one hour up to eight hours in a number of different indoor environments. The objective of the Guideline is to ensure that infected persons such as the one illustrated in the calendar infect less than one person on average over the entire period they are infectious.

**Figure 1:**
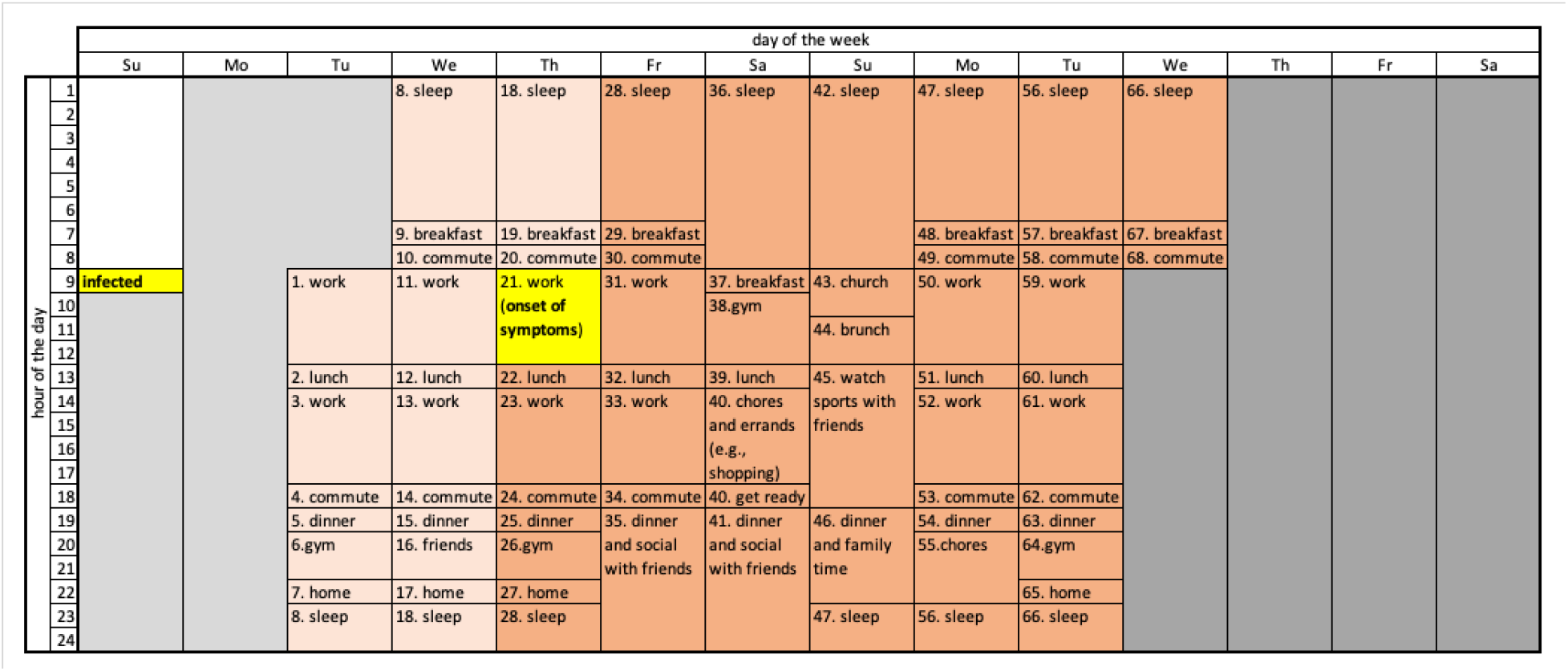
Calendar of typical activities during an infectious period.

The expected number of secondary infections from an infected person is the sum of the expected number of secondary infections from each activity in the sequence over the complete duration that the infected person is infectious.

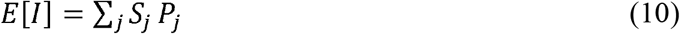

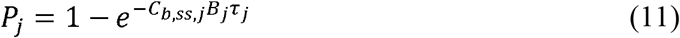

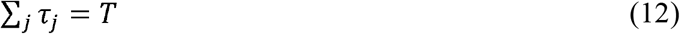

In equations 10-12, j denotes the activity number. In equation 12 and subsequent analysis, T is the mean infectious period. It is equivalent to the inverse of the recovery rate for the SIR model. There are many ways to achieve *E* [*I*]< 1.0 for a sequence of activities. We propose choosing a threshold for the expected number of secondary infections per activity equal to the activity duration divided by the mean infectious period.

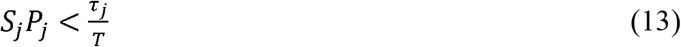

With Equation 11, this becomes the following.

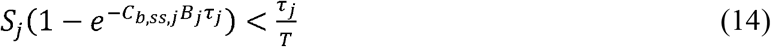

Re-arranging terms we get the following.

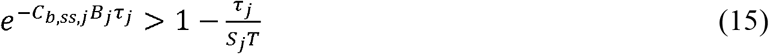

Taking the log of both sides yields the following.

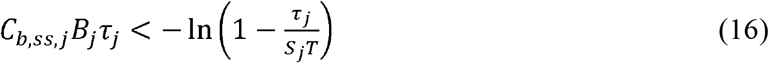

From the Maclaurin series expansion of 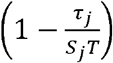 we know that 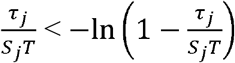.So the following Criterion when applied to every activity ensures condition (13) which ensures that the expected number of secondary infections is less than one.

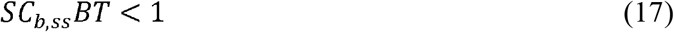

Combining with Equation 5 and re-arranging, the Criterion is the following.

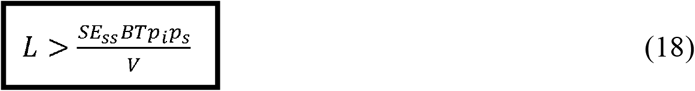

Note that this Criterion is slightly conservative. It yields an expected number of infections over the mean infectious period slightly less than 1.0 even at the threshold. The Criterion is more conservative for mean infectious periods with few activities that last longer and contain fewer susceptible occupants. For example, the expected number of infections for the corner case of a single activity operated at the Criterion threshold lasting T days with just one susceptible occupant 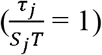 has an expected number of infected people equal to 0.63 rather than 1.0.

We use a default value for COVID-19 mean infectious period, T, of eight days. That accounts for two days of pre-symptomatic infectiousness plus six days of post-symptomatic infectiousness. We choose six post-symptomatic days based on the finding of Cheng et al. (2020) showing that “hospital contacts did not develop infection if their exposure to a case patient started 6 days or more after the case patient’s illness onset.”

We also consider the case where symptomatic people isolate after just two days of symptoms. With 35% of infections being asymptomatic (Mizumoto et al. 2020), this reduces the effective mean infectious period to 5.4 days. For this shortened infectious period to be valid, rapid COVID-19 tests would have to be widely available, and there would have to be strict enforcement of isolation. Neither of these conditions exist in the USA today, nor in most other countries.

While the Guideline has been developed with SARS-CoV-2 in mind, it could also be applied to future epidemics and other pathogens. Doing so would require the use of pathogen-specific characteristics such as the quanta concentration in exhaled breath for people infected with the other pathogen.

The Guideline is equivalent to the indoor safety guideline proposed by Bazant and Bush (2000) with *τ* = T and, *ϵ* = 1. They aimed at determining the safe cumulative time that one could engage in an indoor activity that will keep the probability of becoming infected less than a value *ϵ* ≪ 1. The value of *ϵ* is left up to individuals based on their subjective risk assessment, so the Bazant and Bush guideline is also subjective.

The objective of this Guideline is to control infection at the population level by operating buildings so that the expected number of secondary infections from an infected person is less than 1.0. The Guideline is not designed to control risk for high-risk populations. In situations where the Criterion does not provide a sufficiently low risk it is still possible to increase the loss rate, reduce the occupant density, or use more efficient masks.

Gao et al. (2009) recognized that the infection rate from WR could be substituted into SIR. But they rejected the combined model, stating in their Appendix C that “Applying this model into a community without any change leads to a misuse of two time variables (*τ* and t) in the Wells– Riley equation and SIR model.” Instead, they used computer simulations to illustrate levels of ventilation that would suppress infection rates.

## Applying the Guideline during the COVID-19 Pandemic

The following table shows the inputs used by the Guideline for selected occupancy categories defined in ASHRAE Standard 62.1-2019, plus three different transportation activities and the activity of being at home. The table also shows the pre-pandemic air change rate based on these inputs. For the activities from ASHRAE 62.1-2019, we’ve used the default occupant density. For commercial air travel we used the geometry and occupant density described in You et al. (2019). For subway travel we use the geometry and maximum capacity of New York City subway cars. For rideshare, we use the geometry of a Toyota Prius and the air change rate measured by Ott et al. (2008) with passenger windows closed (with windows open the air change rate is 36/hour). For home we use the midpoint of the closed-windows air change rate in Jimenez (2020) because people are unlikely to keep their windows open during the winter. For the activity column, IS denotes intermediate speech and NNB denotes nose-nose breathing.

The following table shows the Criterion threshold (minimum required values) of *L* for the categories and activities listed in Table 3 for two occupant densities (OD = percent of pre-pandemic floor area per person from Table 3) and three masking conditions assuming all occupants are susceptible. Vaccines and natural immunity reduce the number of susceptible occupants involved in an activity. This increase in immunity can be represented as a reduction in the (susceptible) occupant density for application of the Guideline.

**Table 1:** Efficiency and penetration ratios of masks

| Mask type | $\eta_i$ | $\eta_s$ | $p_i$ | $p_s$ |
| --- | --- | --- | --- | --- |
| N95, KN95, FF2 | 90% | 90% | 10% | 10% |
| N95 with valve | 0% | 90% | 100% | 10% |
| Surgical mask | 50% | 47% | 50% | 53% |
| Cloth mask | 50% | 30% | 50% | 70% |
| Face shield | 23% | 23% | 77% | 77% |

**Table 2:** Quanta concentration in exhaled breath

| Expiratory type | Ci, quanta/m<sup>3</sup> |
| --- | --- |
| Voiced “aah” | 970 |
| Loud speech | 142 |
| Intermediate speech | 72 |
| Quiet speech | 29 |
| Nose-nose breathing | 8.8 |

**Table 3:**
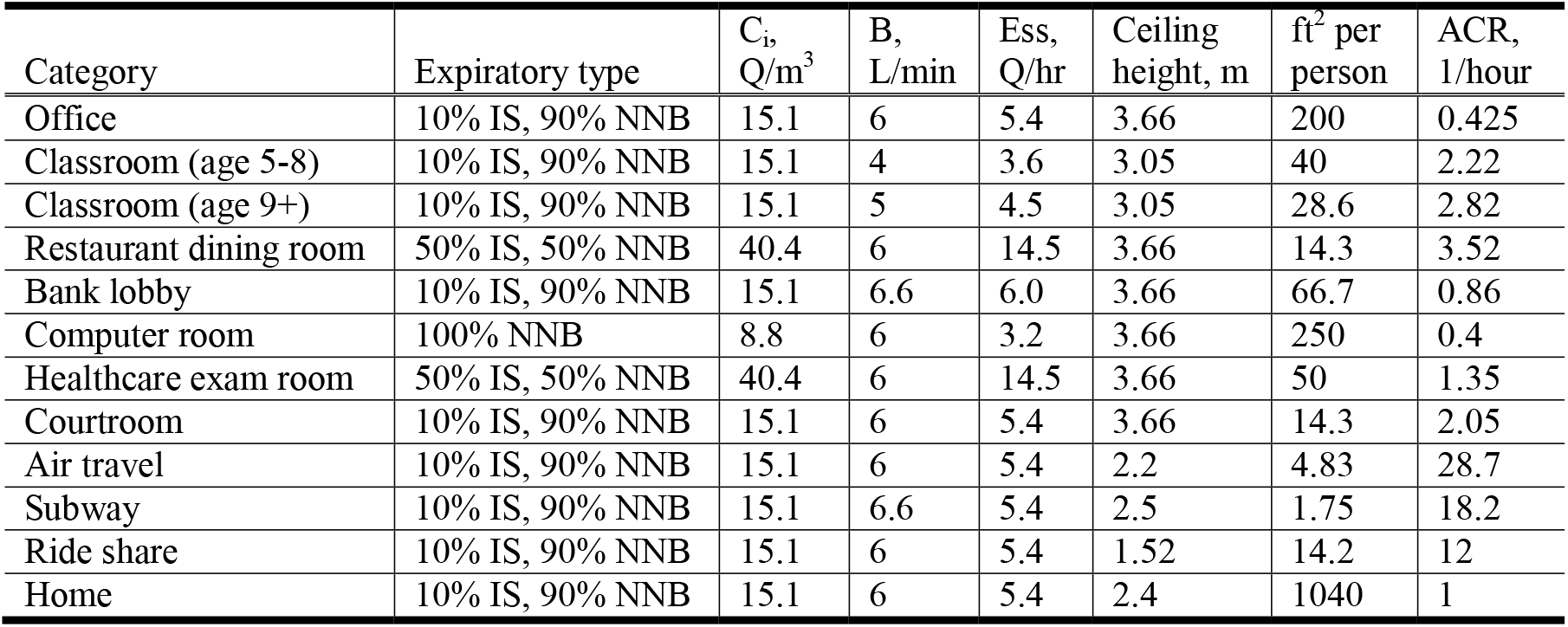
Input conditions and pre-pandemic air change rate for selected activities

The following table shows the Criterion threshold values for a mean infectious period of 5.4 days. To achieve this shorter infectious period would require widespread availability of rapid tests and isolation for all people who test positive. Rapid tests are not yet widely available today, and there is no mechanism to enforce isolation compliance in the USA, nor in many other countries.

## Comparison with Personal Risk Calculations

For a given set of people in a place performing an activity for a duration *τ*, we can compute the Wells-Riley probability when the Criterion is just met 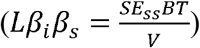. This probability is 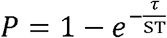. As the number of occupants increases, the WR probability at the Criterion threshold decreases. And as the duration of the activity increases, the WR probability at the Criterion threshold increases.

The following table shows the Wells-Riley probabilities for the occupancy categories of Table 4 assuming all occupants are susceptible and using pre-pandemic occupant densities. For the healthcare exam room, we assume an air-handling unit that serves five exam rooms. For the air travel case, the calculations are based on the seven-row section analyzed by You et al. (2019). For the home, we assume the average size and number of occupants for single-family homes in the USA. Most of the probabilities in Table 6 are very low. They need to be very low so that the expected number of secondary infections over the sequence of activities that an infected person performs over the entire duration of their infection is 1.0 (or less).

**Table 4:** Guideline minimum values of *L* for T=8 days

| Category | No masks |  | Cloth masks |  | N95/KN95 |  |
| --- | --- | --- | --- | --- | --- | --- |
|  | 50% OD | 100% OD | 50% OD | 100% OD | 50% OD | 100% OD |
| Bank lobby | 9.51 | 19.6 | 3.33 | 6.85 | 0.095 | 0.196 |
| Classroom (age 5-8) | 6.57 | 13.9 | 2.3 | 4.85 | 0.066 | 0.139 |
| Classroom (age 9+) | 15.6 | 31.1 | 5.44 | 10.9 | 0.156 | 0.311 |
| Computer room | 1.26 | 2.51 | 0.44 | 0.88 | 0.013 | 0.025 |
| Courtroom | 38.1 | 76.3 | 13.3 | 26.7 | 0.381 | 0.762 |
| Healthcare exam room | N/A | 29.5 | N/A | 10.3 | N/A | 0.295 |
| Office | 2.77 | 5.54 | 0.97 | 1.94 | 0.027 | 0.055 |
| Restaurant dining room | 101 | 204 | N/A | N/A | N/A | N/A |
| Air travel | 181 | 370 | 63.3 | 129 | 1.81 | 3.7 |
| Subway | 554 | 1113 | 194 | 390 | 5.54 | 11.1 |
| Ride share | N/A | 137 | N/A | 48.1 | N/A | 1.37 |
| Home | N/A | 1.46 | N/A | 0.512 | N/A | 0.015 |

**Table 5:** Guideline minimum values of *L* for T=5.4 days

| Category | No masks |  | Cloth Masks |  | N95/KN95 |  |
| --- | --- | --- | --- | --- | --- | --- |
|  | 50% OD | 100% OD | 50% OD | 100% OD | 50% OD | 100% OD |
| Bank lobby | 6.42 | 13.2 | 2.25 | 4.62 | 0.064 | 0.132 |
| Classroom (age 5-8) | 4.43 | 9.38 | 1.55 | 3.27 | 0.045 | 0.094 |
| Classroom (age 9+) | 10.5 | 21.0 | 3.67 | 7.36 | 0.105 | 0.21 |
| Computer room | 0.85 | 1.69 | 0.30 | 0.59 | 0.009 | 0.017 |
| Courtroom | 25.7 | 51.5 | 8.98 | 18.0 | 0.257 | 0.514 |
| Healthcare exam room | N/A | 19.9 | N/A | 6.95 | N/A | 0.199 |
| Office | 1.87 | 3.74 | 0.655 | 1.31 | 0.018 | 0.037 |
| Restaurant dining room | 68.2 | 138 | N/A | N/A | N/A | N/A |
| Air travel | 122 | 250 | 42.7 | 87.1 | 1.22 | 2.50 |
| Ride share | N/A | 92.5 | N/A | 32.5 | N/A | 0.925 |
| Subway | 374 | 751 | 131 | 263 | 3.74 | 7.49 |
| Home | N/A | 0.99 | N/A | 0.344 | N/A | 0.010 |

**Table 6:** Wells-Riley probabilities for Criterion thresholds of Table 4 (OD=100%)

| Category | Duration, hours | # susceptible occupants | Floor area, m<sup>2</sup> | Wells-Riley Probability |
| --- | --- | --- | --- | --- |
| Bank lobby | 0.5 | 37 | 235 | 0.00704% |
| Classroom (age 5-8) | 3 | 19 | 75 | 0.082% |
| Classroom (age 9+) | 6 | 24 | 66 | 0.13% |
| Computer room | 4 | 42 | 1000 | 0.0495% |
| Courtroom | 6 | 104 | 140 | 0.03% |
| Healthcare exam room | 0.5 | 9 | 46 | 0.0289% |
| Office | 8 | 107 | 2000 | 0.039% |
| Restaurant dining room | 2 | 99 | 133 | 0.0105% |
| Air travel | 2.5 | 41 | 18.9 | 0.0318% |
| Ride share | 0.33 | 1 | 1.8 | 0.172% |
| Subway | 0.5 | 251 | 41 | 0.00104% |
| Home | 12 | 2 | 214 | 3.08% |

**Table 7:** Susceptible fraction for pre-pandemic requirements to exceed the Criterion threshold

| Category | No mask | Cloth mask |
| --- | --- | --- |
| Bank lobby | 8.8% | 25% |
| Classroom (age 5-8) | 21% | 60% |
| Classroom (age 9+) | 11% | 30% |
| Computer room | 56% | 100% |
| Courtroom | 3.3% | 9.6% |
| Healthcare exam room | 7% | 20% |
| Office | 26% | 73% |
| Restaurant dining room | 1.8% | N/A |
| Air travel | 5.9% | 17% |
| Subway | 1.3% | 3.7% |
| Ride share | 8.1% | 50% |
| Home | 100% | 100% |

## Emerging from an Epidemic

The loss rate thresholds in Table 4 are significantly higher than the minimum air change rates in Table 3 in order to bring an epidemic under control. But eventually epidemics die out, which begs the question “what does the Guideline look like after the pandemic?” The threshold of the Criterion is dependent on the number of susceptible occupants involved in an indoor activity. Through natural infection and vaccines, the number of susceptible occupants decreases over time even if the number of occupants remains the same every day. So over time, the Criterion threshold becomes lower and lower, in some cases dropping below loss rates corresponding to pre-pandemic ventilation, filtration, inactivation and settling.

The following table shows the fraction of susceptible occupants where pre-pandemic air change rates combined with clean air delivery from MERV8 filters at a heating airflow rate of 5.5 cubic meters per hour per square meter of floor area (0.3 CFM per square foot) and a settling rate of 1.0/hour just equals the Criterion threshold when the occupant density is 100%.

For many of the categories, the susceptible fraction at the Criterion threshold is very low. The susceptible fraction that could keep the loss rate above the Criterion threshold could be higher if higher levels of filtration or outdoor air ventilation were required or if humidification were required. It may be necessary to make permanent changes to codes and standards for minimum ventilation and filtration.

## Summary

The Guideline fulfills a pressing need for a standard way of operating buildings during the COVID-19 pandemic that is scientifically objective, prescriptive, yet flexible, and simple enough to be implemented easily. The Guideline is built on three well-known mathematical models for pathogen accumulation, infection, and population dynamics. If implemented broadly, it would result in the desirable outcome of a population-level reproduction number less than 1.0, driving infections down and helping to extinguish the pandemic. The Guideline’s Criterion can be computed and tabulated for use in conjunction with existing air quality codes and standards. The Guideline could be used to make permanent changes to codes and standards for minimum ventilation and filtration.

## Data Availability

The paper does not make use of external data sets.

## Acknowledgements

The author thanks his colleagues who reviewed the paper and suggested improvements, including Richard Falk, Laurin Herr, Chris Kryzan, Jim Rynne, and Bob Thronson. The author also thanks Pawel Wargocki, Jarek Kurnitski, and Tim Salsbury for their review and feedback.

## Notation

| Symbol | Meaning |
| --- | --- |
| $b$ | Effective contact rate for SIR model |
| $B$ | Breathing rate (e.g., liters/minute) |
| $\beta$ | Filtration beta factor (ratio of contaminant upstream vs downstream of filter) |
| $\beta_i$ | Beta factor of mask worn by infected occupant |
| $\beta_s$ | Beta factor of mask worn by susceptible occupant |
| $C_a$ | Concentration of indoor air (e.g., quanta/m<sup>3</sup>) |
| $\dot{C}_a$ | Time rate of change of the concentration of indoor air |
| $C_{b,ss}$ | Steady state pathogen concentration inhaled by a susceptible occupant |
| $C_i$ | Quanta concentration in the exhaled breath of an infected person |
| $E$ | Emission rate (e.g., quanta/hour) |
| $E_{ss}$ | Steady-state emission rate |
| $F_{CADR}$ | Clean air delivery rate of an in-room air purifier |
| $F_{DOAS}$ | Outdoor airflow rate of a dedicated outdoor air system |
| $F_s$ | Supply airflow rate |
| $\Phi$ | Outdoor air fraction |
| $H$ | Height of indoor space |
| $I$ | Number of infected people |
| $k$ | Inactivation rate |
| $\kappa_f$ | Forced inactivation rate from devices in air-handling equipment |
| $L$ | Combined loss rate (e.g., 1/hour) |
| $M$ | Mechanical air change rate |
| $\nu$ | Recovery rate |
| $N$ | Number of people in a population or an activity |
| $Q$ | Quanta |
| $R$ | Number of recovered or deceased people in a population |
| $S$ | Number of susceptible people in a population or an activity |
| $T$ | Time period that an infected person is infectious |
| $v$ | Settling velocity |
| $V$ | Volume of indoor space |

## References

Bazant, MZ and JWM Bush, Beyond Six Feet: A Guideline to Limit Indoor Airborne Transmission of COVID-19, medRxiv preprint https://doi.org/10.1101/2020.08.26.20182824.

Buonanno, G., Stabile, L. & Morawska, L. Estimation of airborne viral emission: quanta emission rate of SARS-CoV-2 for infection risk assessment. Environment International 141, 105794 (2020), https://doi.org/10.1016/j.envint.2020.105794.

CDC (2020) https://www.cdc.gov/coronavirus/2019-ncov/more/scientific-brief-sars-cov-2.html.

Cheng HW, Jian SW, Liu DP, Ng TC, Huang WT, Lin HH, et al. Contact Tracing Assessment of COVID-19 Transmission Dynamics in Taiwan and Risk at Different Exposure Periods Before and After Symptom Onset. JAMA Intern Med 2020 May 1; doi:10.1001/jamainternmed.2020.2020.

DHS (2020) https://www.dhs.gov/science-and-technology/sars-airborne-calculator.

Gao X, Li Y, and Leung GM, Ventilation Control of Indoor Transmission of Airborne Diseases in an Urban Community, Indoor Built Environ 2009;18;3:205–218, DOI: 10.1177/1420326X09104141.

Jimenez, JL (2020), https://docs.google.com/spreadsheets/d/16K1OQkLD4BjgBdO8ePj6ytf-RpPMlJ6aXFg3PrIQBbQ/htmlview?pru=AAABdZlTFtc*hiUcDrNUq7CsH3X_D5yYvg#gid=519189277.

Lewis, E. R. (2008), An examination of Kohler theory resulting in an accurate expression for the equilibrium radius ratio of a hygroscopic aerosol particle valid up to and including relative humidity 100%, J. Geophys. Res., 113, D03205, doi:10.1029/2007JD008590.

Miller SL, Nazaroff WW, Jimenez JL, et al. Transmission of SARS-CoV-2 by inhalation of respiratory aerosol in the Skagit Valley Chorale superspreading event. Indoor Air. 2020;00:1–10. https://doi.org/10.1111/ina.12751.

Mizumoto K, Kagaya K, Zarebski A, Chowell G. Estimating the asymptomatic proportion of coronavirus disease 2019 (COVID-19) cases on board the Diamond Princess cruise ship, Yokohama, Japan, 2020. Euro Surveill. 2020;25(10):pii=2000180. https://doi.org/10.2807/1560-7917. ES.2020.25.10.2000180.

Oberg, T. and Brosseau, LM. 2008. Surgical mask filter and fit performance. American journal of infection control 36, 276–282, doi:10.1016/j.ajic.2007.07.008.

You, R., Lin, C.-H., Wei, D., and Chen, Q. 2019. “Evaluating the commercial airliner cabin environment with different air distribution systems,” Indoor Air, 29:840–853.

